# Evaluating the Diagnostic Performance of Large Language Models in Identifying Complex Multisystemic Syndromes: A Comparative Study with Radiology Residents

**DOI:** 10.1101/2024.06.05.24308335

**Authors:** Hagar Olshaker, Dana Brin, Elinor Kalderon, Matan Kraus, Eliahu Konen, Eyal Klang

**Author notes:** **Corresponding authors:** Hagar Olshaker,Eyal Klang. Financial disclosure – none.

## Abstract

**Aim:** This study evaluates the diagnostic capabilities of large language models (LLMs) in interpreting imaging patterns, focusing on their utility as a resource for radiology residents. We compare the diagnostic performance of OpenAI’s GPT-3.5, GPT-4, and Google’s Gemini Pro against radiology residents in identifying complex, multisystemic syndromes with an increased risk of cancer.

**Methods:** We assessed diagnostic accuracy using textual descriptions of radiological findings from 60 diseases selected from The Familial Cancer Database. Participants included three LLMs and three radiology residents. Diagnostic responses were scored on accuracy and first choice correctness. Experiments with AI models were conducted using default API settings.

**Results:** GPT-4 achieved the highest diagnostic accuracy (63%) and first choice accuracy (40%), significantly outperforming the radiology residents whose accuracy ranged from 22% to 43%. The overall average accuracy for AI models was 49.3%, compared to 29.0% for residents. Error analysis revealed that while some diseases were universally recognized, others highlighted diagnostic challenges across both human and AI participants.

**Conclusion:** GPT-4 outperforms radiology residents in diagnosing complex, infrequent multisystemic diseases. These findings suggest potential benefits of integrating AI tools to improve diagnostic accuracy for rare conditions and imply a need for revisions in medical training to incorporate AI competencies, enhancing diagnostic processes and resident education in radiology.

## Introduction

The rapid growth in radiological data presents significant challenges for radiologists(1). This surge necessitates that radiologists enhance their productivity.

One of the challenges faced by radiologists is the detection of multisystemic syndromes, which are inherently complex due to the involvement of various body systems (2–6). This task is particularly critical when the syndrome predisposes an individual to cancer. In such cases, rapid and accurate detection is crucial for implementing treatment strategies. The complexity of these conditions requires radiologists to possess a high level of expertise to ensure precise diagnosis(7).

Artificial Intelligence (AI) is increasingly recognized for its potential to address such demanding tasks. AI refers to a field of computer science dedicated to creating systems capable of performing tasks that typically require human intelligence (8). In radiology, AI has been shown to be most promising as a means of improving image acquisition, reconstruction and interpertation(9). Large Language Models (LLMs) are advanced AI systems designed to process, understand, and generate human-like text, enabling them to perform a variety of language-based tasks.

One of the most significant contributions of AI to radiology so far is its ability to detect findings from imaging tests using image recognition(10). However, with the rise of LLMs, it is becoming clear that AI applications are not limited to imaging-related tasks in radiology (11,12).

Our study aims to evaluate the diagnostic capabilities of LLMs in interpreting imaging patterns, focusing on their utility as a resource for radiology residents. We compare the diagnostic performances of three leading LLMs—OpenAI’s GPT-3.5, GPT-4, and Google’s Gemini Pro— against those of radiology residents in identifying complex, often multisystemic syndromes that carry an increased risk of cancer.

## Methods

### Study Design and Participants

This study evaluated the diagnostic accuracy of AI models and radiology residents in identifying complex, often multisystemic syndromes that, while not primarily oncological, carry an elevated risk of cancer development. The diseases were selected from The Familial Cancer Database (FaCD).

Participants included three LLMs (GPT-3.5, GPT-4, Gemini Pro) and three radiology residents: one with 1.5 years in residency and two with 2.5 years in residency. Participants were shown textual descriptions of radiological findings from patient cases obtained from the collaborative website Radiopaedia.org.

### Data Collection

Diagnostic responses for 60 rare diseases were collected from each participant, using the standardized prompt: “Please give a differential diagnosis that explains the following imaging findings:…“ to ensure uniformity. At the beginning of each description, the modality type was specified, using only CT and MRI, as these modalities are most familiar to our institution’s residents, ensuring no technical misunderstandings influenced the study outcomes. All participants were blinded to each other’s responses. The responses were scored as 0 (incorrect diagnosis), 1 (correct diagnosis as first choice), or 2 (correct diagnosis but not as first choice).

### AI Model Experimentation

The experiments with AI models were conducted using the OpenAI API for GPT-3.5 and GPT-4, and Google’s Bard API for Gemini Pro. Default API hyper-parameters such as models’ temperature were employed for all the models. All experiments were performed using Python version 3.9.

### Statistical Analysis

Diagnostic accuracy was calculated as the proportion of cases where participants correctly diagnosed diseases, with scores of 1 or 2. We employed Tukey’s Honest Significant Difference (HSD) test to compare the mean accuracies of AI models and residents. A p-value of less than 0.05 was considered statistically significant.

An error analysis was also performed, categorizing diseases based on diagnostic accuracy across all participants. Categories included ‘All Participants Right’, ‘Only GPT-4 Right’, and ‘All Participants Wrong’.

## Results

### Accuracy of Diagnoses

In this study, we assessed the accuracy of AI models (GPT-3.5, GPT-4, Gemini Pro) and radiology residents in identifying complex, often multisystemic diseases, that carry an increased risk of cancer development. The results are presented in **Table 1**.

**Table 1:** Accuracy of AI Models and Radiology Residents.

| Participant | Accuracy |
| --- | --- |
| GPT-3.5 | 30/60 (50%) |
| GPT-4 | 38/60 (63%) |
| Gemini Pro | 21/60 (35%) |
| Resident 1 (D) | 13/60 (22%) |
| Resident 2 (M) | 26/60 (43%) |
| Resident 3 (E) | 13/60 (22%) |
| Average (Models) | 29.6/60 (49.3%) |
| Average (Residents) | 17.3/60 (29.0%) |

GPT-4 demonstrated the highest accuracy (63%), while Gemini Pro showed the lowest among the AI models (35%). Residents’ accuracies varied from 22% to 43%. The overall average accuracy for AI models was higher (49.3%) compared to that of the radiology residents (29.0%).

### First Choice Accuracy of Diagnoses

We further evaluated instances where participants correctly identified the diagnosis as their initial choice. The findings are presented in **Table 2**.

**Table 2:** First Choice Accuracy of AI Models and Radiology Residents.

| Participant | Accuracy |
| --- | --- |
| GPT-3.5 | 15/60 (25%) |
| GPT-4 | 24/60 (40%) |
| Gemini Pro | 16/60 (27%) |
| Resident 1 (D) | 12/60 (20%) |
| Resident 2 (M) | 23/60 (38%) |
| Resident 3 (E) | 10/60 (17%) |
| Average (Models) | 18.3/60 (30.5%) |
| Average (Residents) | 15/60 (25.0%) |

GPT-4 displayed the highest first choice accuracy at 40%. The residents first choice accuracies varied between 17-38%. The overall average first choice accuracy for AI models was 30.5%, compared to 25% for the radiology residents.

### Statistical Significance Analysis

In assessing the statistical significance of diagnostic accuracies, distinct patterns emerged among AI models and residents:

### GPT-4 Performance

GPT-4 demonstrated statistically significant superiority over the residents with p-values of <0.001, <0.001, and 0.019, respectively. Additionally, its performance was significantly better than Gemini Pro (p=0.005). However, no statistically significant difference was observed when comparing GPT-4 with GPT-3.5.

### GPT-3.5 Accuracy

GPT-3.5 showed significantly higher accuracy than two of the residents (p<0.001 for both), yet its accuracy did not significantly differ from that of Resident 2 or Gemini Pro.

### Gemini Pro Comparison

Gemini Pro’s performance did not exhibit statistically significant differences compared to the residents.

This highlights GPT-4’s relative effectiveness in this context.

#### Error Analysis

We conducted an error analysis to understand the diagnostic accuracy patterns among AI models and radiology residents. The analysis, detailed in **Table 3**, revealed three distinct categories of diagnostic outcomes.

**Table 3:** Summary of Error Analysis in Disease Diagnosis.

| Category | N (%) | Diseases |
| --- | --- | --- |
| All Participants<br>Right | 4/60<br>(6.7%) | Alpha-1-antitrypsin deficiency<br>Autosomal dominant polycystic kidney disease<br>Diabetes Mellitus (case of emphysematous pyelonephritis)<br>Meningiomatosis |
| Only GPT-4<br>Right | 7/60<br>(11.7%) | Blue rubber bleb nevus syndrome<br>Cherubism |
|  |  | Neurofibromatosis type 1 (NF1)<br>Neurofibromatosis type 2 (NF2)<br>Proteus syndrome<br>Rubinstein-Taybi syndrome<br>Ataxia-telangiectasia<br>Common variable immunodeficiency (CVID) |
| All Participants<br>Wrong | 15/60<br>(25%) | Birt-Hogg-Dubé syndrome<br>Currarino syndrome<br>McCune-Albright syndrome<br>Muenke syndrome<br>Neuronal ceroid lipofuscinosis<br>Down syndrome<br>Fragile X-associated tremor/ataxia syndrome (FXTAS)<br>Gaucher's disease<br>Glycogen storage disease type Ib |
|  |  | Hereditary angioedema (case of small bowel)<br>Incontinentia pigmenti (Bloch-Sulzberger syndrome)<br>Juvenile hyaline fibromatosis<br>Multiple Endocrine Neoplasia Type 1 (MEN1)<br>Chromosome 18q deletion syndrome (18q syndrome) |

Firstly, certain diseases, like Alpha-1-antitrypsin deficiency, were correctly identified by all participants. This consistency suggests a general familiarity with these conditions among both humans and AI models. Such diseases likely have well-defined diagnostic criteria that are widely recognized.

Intriguingly, GPT-4 stood out by correctly diagnosing 7/60 (11.7%) complex diseases, such as Neurofibromatosis types 1 and 2, where other participants faltered. This disparity in diagnostic ability suggests that AI models like GPT-4 can offer valuable insights in situations where traditional medical expertise may be limited.

On the other hand, some diseases presented significant challenges to all participants. For instance, Birt-Hogg-Dubé syndrome and Currarino syndrome were consistently misdiagnosed by everyone. These cases, making up 15/60 (25%) of the cases, signal potential gaps in both medical training and AI development, especially concerning rare or atypical disease presentations.

## Discussion

Our study demonstrates that LLMs, particularly GPT-4, show promise in identifying differential diagnoses in complex syndromes, surpressing the level of several radiology residents.

Many studies have attempted to demonstrate the superior performance of AI software over radiologists in detecting imaging findings. One recently published study examines the performance of AI in interpreting digital mammography for breast cancer screening. The findings showed that AI generally matched or exceeded the performance of radiologists in identifying breast cancer from mammograms(13). Another study explores the effectiveness of an AI algorithm compared to radiology residents in interpreting frontal chest radiographs, with the joint interpretation of three board-certified radiologists set as control. The results were that the AI algorithm and the radiology residents performed similarly in terms of detecting the various findings, however, the AI algorithm showed higher specificity and positive predictive value than the residents (14).

Moreover, many studies were published showing AI software can match the abilities of doctors, particularly residents, in all kinds of tasks, such as in board exams in many fields(15,16) (but not radiology(17)), triage and clinical diagnosis (18,19), and more (20,21). These recent studies show that although AI programs cannot yet replace doctors (22,23), it has a potential to enhance the health care abilities in providing the best medical care.

In our study, we demonstrate that using LLM can help in two ways. First, it can make the diagnostic process more efficient, as identifying rare and multisystem diseases can be difficult and time-consuming due to the variable and multisystemic manifestation. Second, it can help residents which can benefit from a tool that can easily answer complex questions. In light of these findings and others(24), it’s worth considering a reevaluation and update of medical residents’ training such as integration of AI tools into the diagnostic process, at least in some medical fields(25). This emerging capability represents a paradigm shift in medical education and practice(26), and as such, it underscores the importance of equipping healthcare professionals with the skills and knowledge to effectively leverage these technologies in clinical settings, balancing traditional diagnostic methods with AI-driven insights.

Our study has limitations, including a small sample size and participants from one institution, which may affect the generalizability of the results. Using only textual descriptions from a single source for radiological findings might not reflect the complexity of real-world diagnostics. Additionally, the AI models were used with default settings without customization for specific radiological tasks. Further exploration of AI’s utility should consider clinical applicability and diagnostic time to fully assess its integration into medical practice. Finally, we did not explore techniques such as retrieval augmented generation (RAG) or fine-tuning.

In Conclusion, our study demonstrates that GPT-4 outperforms radiology residents in diagnosing complex, infrequent multisystemic diseases. This finding suggests that integrating advanced AI tools could improve diagnostic accuracy for such conditions. It also indicates a need to revisit and potentially revise the training of radiology residents to include AI competencies.

## Data Availability

All data produced in the present study are available upon reasonable request to the authors

## Acknowledgments

none

